# A Survey of Open Science Attitudes and Behaviors among US Pharmacy Faculty

**DOI:** 10.1101/2024.06.20.24309260

**Authors:** Spencer E. Harpe

**Affiliations:** Professor of Pharmacy Administration Midwestern University College of Pharmacy, (Downers Grove, IL Campus) 555 31st Street, Downers Grove, IL 60515 USA

**Keywords:** Open science, Preregistration, Preprints, Open access, Data sharing, Meta-research

## Abstract

Objectives: To describe the current attitudes, behaviors, and perceived disciplinary norms related to open science practices among full-time pharmacy faculty in the US and to examine differences in attitudes and behaviors across pharmacy disciplines.

Methods: In this cross-sectional study, the Center for Open Science’s Open Scholarship Survey modules on data sharing, code sharing, materials sharing, preregistration, preprints, and open access publishing were administered to a random sample of 3,200 faculty from the AACP Roster of Pharmacy Faculty as of February 2022. Individuals with at least a 0.8 full-time equivalent faculty appointment in pharmacy practice or one of the pharmaceutical sciences were eligible to participate.

Results: Responses were obtained from 663 faculty (502 complete; 161 partial). The most positive attitudes were for open access publishing (overall mean [SD]: 4.1 [0.9]) with the lowest attitudes for study preregistration (3.2 [0.9]) and posting preprints (3.1 [1.1]). Statistically significant differences in attitudes across pharmacy disciplines were identified for data sharing, code sharing, and study preregistration. The most commonly reported open science practice was open access publishing (mean [SD], 27.7% [29.1%]). Study preregistration was the least common (mean [SD], 1.7% [7.0%]). After accounting for respondent and institutional characteristics, differences in open science behaviors were noted across pharmacy disciplines.

Conclusion: This study provides a baseline assessment of faculty attitudes towards and engagement in open science practices among US pharmacy faculty. Given the relatively low frequency with which open science practices were reported, there is considerable room for improvement in the uptake of open science practices.

## 1. Introduction

According to the United Nations Educational, Scientific, and Cultural Organization (UNESCO), open science is “an inclusive construct that combines various movements and practices aiming to make multilingual scientific knowledge openly available, accessible and reusable for everyone, to increase scientific collaborations and sharing of information for the benefits of science and society, and to open the processes of scientific knowledge creation, evaluation and communication to societal actors beyond the traditional scientific community.”^1^ UNESCO’s recommendations required open science infrastructures, open engagement of societal actors, open dialog with other knowledge systems, and open scientific knowledge.^1^ The US National Academies of Science, Engineering, and Medicine have emphasized that open science involves not only the *products* of science (i.e., evidence generated through research), but also the *processes* of science in all their various forms.^2^

Two commonly cited benefits of open science relate to increased availability and transparency of research to other researchers and society at large.^3^ By increasing transparency, open science may increase trust in the processes and products of science.^4,5^ With the high estimates of research waste,^6,7^ open science may improve efficiency and reduce waste by supporting collaboration amongst researchers and promoting the reuse of existing data.^2,3,8,9^ Engaging in open science provides benefits to individual researchers, such as increased citations, employability, and research collaborations.^4,10,11^ From a holistic perspective, open science has been proposed as a means to improve equity in the scientific enterprise for researchers, research participants, and research consumers alike.^1–3,12^

Open science is not without potential challenges or disadvantages. There remain notable costs for open access publishing.^13^ Some in the scientific community have raised the concerns surrounding the privacy of research participants or communities^14^ and even potential security threats^15^ associated with sharing research data and materials. Despite the intentions to promote research equity, open science may inadvertently increase inequities.^16,17^

Recent research has sought to examine attitudes and behaviors related to various open science practices. The State of Open Data survey, conducted annually since 2016, has shown increasingly positive views about data sharing as a requirement for funding, especially among early career researchers.^18^ Previous research on open science practices has included a broad range of disciplines across the physical, life, social, and health sciences.^9,19–21^ Although health sciences and health professions have been included in previous studies, pharmacy was not able to be viewed separately. This study provides a baseline of open science attitudes and behaviors among US pharmacy faculty allowing for the detection of changes in the future.

The objectives of this study were (1) to describe the current attitudes, behaviors, and perceived disciplinary norms related to selected open science practices among full-time pharmacy faculty in the US and (2) to examine potential differences in current attitudes and behaviors across pharmacy disciplines.

## 2. Methods

### 2.1. Study Design and Sample

This cross-sectional study included a random sample of faculty from the AACP Roster of Pharmacy Faculty as of February 2022. This roster includes an estimated 6,500 faculty across pharmacy practice and the pharmaceutical sciences. Participants were eligible for inclusion if they had at least a 0.8 full-time equivalent faculty appointment in pharmacy practice or one of the pharmaceutical sciences at a school or college of pharmacy in the US. There were no other restrictions placed on type of faculty position (clinical, teaching, research, tenure-track, etc.). Chief executive officer (CEO) deans listed in the Directory of Professional Programs of Colleges and Schools of Pharmacy maintained by the Accreditation Council for Pharmacy Education in March 2022 were excluded from the roster prior to sampling faculty for questionnaire administration; however, respondents who identified as CEO deans were retained in the data. Based on a total population size of 6,500, a 95% confidence level, and a 3% margin of error, a total sample of 363 was needed for accurate and reliable results. Using a conservative estimated response rate of 15%, which is consistent with previous online surveys of pharmacy faculty,^22,23^ a total sample of 3,200 was drawn randomly from the AACP faculty roster.

### 2.2. Study Questionnaire and Administration

The study questionnaire was based on the Open Scholarship Survey (OSS), a modular survey developed by the Center for Open Science.^24^ The modules on data sharing, code sharing, materials sharing, preregistration, preprints, and open access publishing were used for this study. Questions within each module related to respondents’ attitudes, behaviors, and perceived disciplinary norms (i.e., the percentage of other faculty within their discipline who have engaged in various open science activities and the distribution of attitudes within their discipline towards various open science activities). As part of a larger study, the full study questionnaire included a total of 217 to 220 questions depending on the responses selected as skip logic used as part of the questionnaire flow. The questionnaire was broken into 4 sections: open science attitudes and behaviors, open science incentives and barriers, open science knowledge and future use, and demographics. Only the questionnaire sections on open science attitudes and behaviors and demographics are reported here.

Recruitment took place in 8 waves of 400 randomly selected faculty. Each wave lasted 2 weeks. As the current best, evidence-based practice for conducting survey research, Dillman’s tailored design method^25^ was used as a framework for survey administration. The online questionnaire was open for 2 weeks within each recruitment wave. Each sampled faculty member received an introductory email that included a personalized link to the online questionnaire to prevent duplicate responses. Reminder emails were sent to non-respondents 3, 6, and 10 business days after the introductory email. Questionnaire administration occurred between April 26 and June 26, 2022, using the Qualtrics XM online survey platform (Qualtrics LLC; Provo, UT). The order of the OSS modules was randomized for each respondent to avoid any potential ordering effects. All other questionnaire sections were presented in the same order across respondents. Individuals who completed the survey were offered a $10 gift card to a selection of nationwide retailers. Identifiable information (names, email addresses, IP addresses, etc.) was not collected as part of questionnaire administration. Mailing information for gift cards was collected separately and not linked to any questionnaire responses.

To approximate attentiveness when completing the questionnaire, time spent on each questionnaire page was measured through embedded timer questions. Respondents spending >1 standard deviation below the mean response time across all respondents for the questionnaire page were excluded. If a respondent exceeded the threshold for ≥80% of survey pages, all their responses were excluded.

### 2.3. Study Variables

The OSS modules collected information on attitudes, behaviors, and perceived disciplinary norms related to open science attitudes and behaviors. Attitudes towards open science activities were obtained from the relevant OSS modules using the following response options: Very much against, Against, Neither in favor nor against, In favor, Very much in favor, and No opinion. For open science behaviors, respondents provided the percentage of their scholarly works that involved the 6 open science practices examined in this study. Disciplinary norms for attitudes were assessed by asking respondents to estimate the approximate percentage of faculty within their discipline who fell into each attitude category (Very much against, Against, etc.) for each open science practice. For behavioral norms, respondents estimated the percentage of faculty within their discipline who had engaged in each open science practice.

Individual characteristics (time since highest degree, gender, pharmacy discipline, administrative position, etc.) were obtained from each respondent. Institutional characteristics were obtained from publicly available resources. The standardized set of pharmacy disciplines maintained by AACP was used for this study. Research funding from the 2021-2022 fiscal year research was obtained from AACP Funded Research Grant Institutional Tables.^26^

### 2.4. Statistical Analysis

Representativeness of the study sample was assessed by comparing respondent characteristics to the 2021-2022 academic year AACP Profile of Pharmacy Faculty.^27^ The potential for non-response bias was assessed by comparing respondent characteristics and attitudes and behaviors for early and late responders within waves (individuals responding in the first 3 days of the wave vs. individuals responding on or after the last reminder within the wave) and across the data collection period (Waves 1 and 2 vs. Waves 7 and 8).

Current attitudes were described using means and standard deviations and percentage of responses in each response category. Behaviors were described as the percentage of faculty who had any scholarly works using the open science practice. For the comparison of attitudes and perceived attitudinal norms, the “Very much in favor” and “In favor” responses were combined to describe the total percentage of respondents with favorable attitudes towards each open science practice. Differences in attitudes towards open science practices across pharmacy disciplines were analyzed via ordinary least squares regression with response on the attitude item for each open science practice as the outcome variable and pharmacy discipline as the predictor variable.

For open science behaviors, the proportion of scholarly outputs using the open science practice of interest was the outcome variable for a generalized linear model (GLM) with a log link and binomial distribution. For open science behaviors, three models were constructed: Model 1 (pharmacy discipline only), Model 2 (pharmacy discipline and respondent characteristics), and Model 3 (pharmacy discipline, respondent characteristics, and institutional characteristics). Robust standard errors were estimated to account for clustering of respondents within institutions for all models.

Statistical analyses were conducted in R version 4.4.0 (R Foundation, Vienna, Austria) with p < 0.05 denoting statistical significance. As a pre-specified analysis, comparisons of attitudes across pharmacy discipline were adjusted for multiple comparisons using the Bonferroni method. No additional adjustments were made for the multiple testing across the analyses conducted for this study.^28^ Disciplinary norms for attitudes and behaviors were compared to respondents’ attitudes and behaviors without formal statistical analysis. Respondents not providing their pharmacy discipline, indicating “No opinion” for attitudes towards an open science practice, or not providing any response for attitudes or behavior for an open science practice were excluded from statistical analyses; however, all eligible respondents were included when reporting respondent demographics.

### 2.5. Ethical Approval and Study Registration

This study was reviewed by the Midwestern University Illinois Campus Institutional Review Board and determined to qualify for an exemption (File Number 22011). The study protocol was registered at Open Science Framework Registries (https://osf.io/68rvy) prior to data collection.

## 3. Results

### 3.1. Response Rate and Missing Data

Of the 3200 emails sent, 49 were unable to be delivered. Of those delivered, 116 individuals opted out of participation, and 59 did not meet the eligibility criteria upon completing the screening questions.

Complete responses were obtained from 502 faculty with an additional 161 providing partial responses yielding the following response rates using the American Association of Public Opinion Research^29^ definitions: 16.0% (RR1, complete responses only) and 21.2% (RR2, complete and partial responses). Given the availability of additional financial resources after study initiation, recruitment was not concluded once the original sample size requirement was met.

No respondents were excluded because of insufficient attentiveness. Missing attitudes ranged from 103 for open access publishing to 219 for sharing code. Missing frequency of open science practices ranged from 134 for open access publishing to 177 for sharing code. By randomizing the order of the open science attitude and behavior sections, missingness was assumed to be at random for those variables.

Pharmacy discipline was not provided by 175 respondents. The effective sample sizes are provided in the appropriate tables for each analysis.

### 3.2. Sample Representativeness and Non-Response Bias

Most characteristics of the respondents were within a 5 percentage-point difference of the pharmacy faculty profile with a few exceptions. Faculty from the economic, social, and administrative sciences were more common in the sample (15.9%) than in the faculty profile (7.2%). Faculty at the rank of assistant professor were somewhat underrepresented in the sample (24.6%) compared to the faculty profile (31.7%). There were more tenured faculty responding (36.7%) and fewer tenure-eligible faculty (14.4%) compared to the faculty profile (14.3% and 32.3%, respectively). The full comparisons for representativeness can be found in Supplemental File 1.

Based on the comparisons of early and late responders, there was little concern for non-response bias. Respondent characteristics were similar across early and late respondents both within waves and across the data collection period (Waves 1/2 vs. Waves 7/8). No statistically significant differences were noted in the comparisons and the absolute differences were generally within 5 percentage points. When comparing open science attitudes and behaviors, there were some differences to note. For within-wave comparisons, the mean attitudes were not statistically significantly different; however, late respondents appeared to be more divided on attitudes toward data sharing (i.e., fewer “neither in favor nor against” responses) and were more positive with respect to material sharing. For comparisons across the data collection period, late wave respondents appeared to have slightly less positive attitudes surrounding code sharing than early wave respondents. Although late wave respondents had a statistically higher mean percentage of scholarly works published under open access pathways, there was no difference in the percentage of respondents who had any scholarly work published as open access. The full non-response assessment can be found in Supplemental File 2.

### 3.3. Respondent Characteristics

Respondents were primarily women (53.5%) with White racial identities (72.3%). Over half of the respondents were from the pharmacy practice discipline. Respondents were approximately equally distributed across academic rank with slightly higher representation of associate professors. Time in academia was approximately equally distributed with slightly fewer respondents in the 16- to 20-year category (14.5%). Respondents reported having published a mean (SD) of 27.3 (40.2) studies (median (IQR): 15 (4, 32)). The reported mean (SD) of total publications was 42.9 (58.9) with a median (IQR) of 25 (11, 50). Additional respondent characteristics are provided in Table 1. Based on the distribution of responses, pharmacy disciplines were collapsed into the following groups: pharmaceutical sciences (biological and biomedical sciences; medicinal or pharmaceutical chemistry and pharmacognosy; pharmaceutics; pharmacology and toxicology); economic, social, and administrative sciences (ESAS); pharmacy practice; and clinical sciences (pharmacokinetics, pharmacodynamics, genetics, pharmacotherapeutics, translational research). This was done to streamline analysis and reporting while maintaining groupings that were generally similar in nature with respect to research topics and approaches.

**Table 1:** Respondent Characteristics

|  | No. (%) |
| --- | --- |
| Gender (n = 503)* |  |
| Woman | 270 (53.7%) |
| Man | 206 (41%) |
| Something else (Other, Prefer not to say, etc.) | 29 (5.8%) |
| Race and ethnicity (n = 354)* |  |
| Black or African American | 13 (3.7%) |
| White | 270 (76.3%) |
| Asian | 52 (14.7%) |
| Hispanic or Latinx | 15 (4.2%) |
| Other | 5 (1.4%) |
| Two or more indicated | 8 (2.3%) |
| Unknown or Prefer not to answer | 9 (2.5%) |
| Time in academia (n = 504) |  |
| ≤ 5 years | 97 (19.2%) |
| 6-10 years | 113 (22.4%) |
| 11-15 years | 101 (20.0%) |
| 16-20 years | 73 (14.5%) |
| > 20 years | 120 (23.8%) |
| Degrees earned (n = 513)* |  |
| Bachelor's | 272 (53.0%) |
| Master's (MS, MPH, MBA, etc.) | 145 (28.3%) |
| PharmD | 329 (64.1%) |
| MD or DO | 4 (0.8%) |
| PhD or ScD | 196 (38.2%) |
| Other doctoral degree (DrPH, EdD, etc.) | 7 (1.4%) |
| Currently enrolled in an educational training program (n = 500) | 45 (9.0%) |
| Academic rank (n = 501) |  |
| Professor | 151 (30.1%) |
| Associate Professor | 192 (38.3%) |
| Assistant Professor | 149 (29.7%) |
| Other | 9 (1.8%) |
| Pharmacy discipline (n = 496) |  |
| Biological or biomedical sciences | 37 (7.5%) |
| Medicinal or pharmaceutical chemistry and pharmacognosy | 24 (4.8%) |
| Pharmaceutics | 16 (3.2%) |
| Pharmacology and toxicology | 40 (8.1%) |
| Economic, social, and administrative sciences | 79 (15.9%) |
| Pharmacy practice | 270 (54.4%) |
| Pharmacokinetics, pharmacodynamics, and genetics | 10 (2.0%) |
| Pharmacotherapeutics and translational research | 20 (4.0%) |
| Tenure status (n = 501) |  |
| Tenured | 184 (36.7%) |
| Tenure eligible | 72 (14.4%) |
| Not tenure eligible (institution offers tenure) | 172 (34.3%) |
| Not tenure eligible (institution does not offer tenure) | 73 (14.6%) |

\*Respondents could select more than one option, so percentages may not sum to 100%.

### 3.4. Attitudes Towards Open Science Practices

Attitudes were generally neutral (neither in favor nor against) to positive for all open science practices. The most positive attitudes were related to open access publishing (overall mean [SD]: 4.1 [0.9]) with the lowest attitudes related to study preregistration (3.2 [0.9]) and posting preprints (3.1 [1.1]). The mean (SD) attitudes across pharmacy disciplines are presented in Table 2. Statistically significant differences in attitudes across pharmacy disciplines were identified for data sharing, code sharing, and study preregistration. The largest differences across disciplines were noted for study preregistration where the mean (SD) attitudes for clinical sciences was 3.5 (1.5) compared to 2.9 (1.0) for pharmaceutical sciences. There were only 2 instances where mean responses were “against” an open science practice: study preregistration among pharmaceutical sciences faculty and posting preprints for clinical sciences faculty.

**Table 2:**
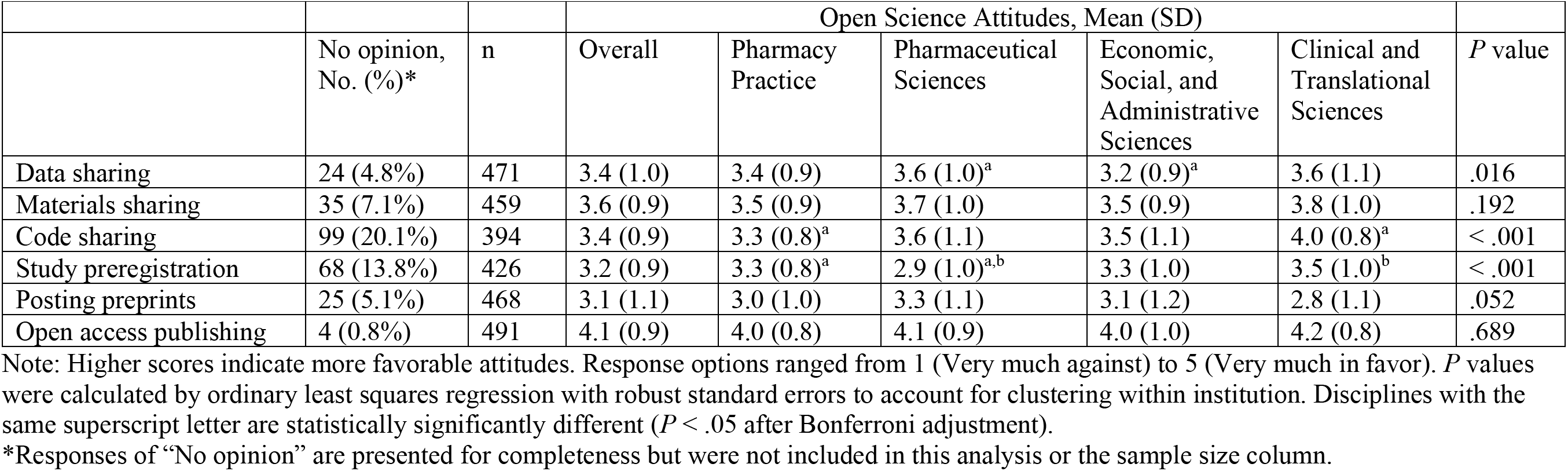
Attitudes towards Open Science Practices by Pharmacy Discipline

|  |  |  | Open Science Attitudes, Mean (SD) |  |  |  |  |  |
| --- | --- | --- | --- | --- | --- | --- | --- | --- |
|  | No opinion,<br>No. (%)* | n | Overall | Pharmacy<br>Practice | Pharmaceutical<br>Sciences | Economic,<br>Social, and<br>Administrative<br>Sciences | Clinical and<br>Translational<br>Sciences | *P* value |
| Data sharing | 24 (4.8%) | 471 | 3.4 (1.0) | 3.4 (0.9) | 3.6 (1.0)<sup>a</sup> | 3.2 (0.9)<sup>a</sup> | 3.6 (1.1) | .016 |
| Materials sharing | 35 (7.1%) | 459 | 3.6 (0.9) | 3.5 (0.9) | 3.7 (1.0) | 3.5 (0.9) | 3.8 (1.0) | .192 |
| Code sharing | 99 (20.1%) | 394 | 3.4 (0.9) | 3.3 (0.8)<sup>a</sup> | 3.6 (1.1) | 3.5 (1.1) | 4.0 (0.8)<sup>a</sup> | < .001 |
| Study preregistration | 68 (13.8%) | 426 | 3.2 (0.9) | 3.3 (0.8)<sup>a</sup> | 2.9 (1.0)<sup>a,b</sup> | 3.3 (1.0) | 3.5 (1.0)<sup>b</sup> | < .001 |
| Posting preprints | 25 (5.1%) | 468 | 3.1 (1.1) | 3.0 (1.0) | 3.3 (1.1) | 3.1 (1.2) | 2.8 (1.1) | .052 |
| Open access publishing | 4 (0.8%) | 491 | 4.1 (0.9) | 4.0 (0.8) | 4.1 (0.9) | 4.0 (1.0) | 4.2 (0.8) | .689 |

Note: Higher scores indicate more favorable attitudes. Response options ranged from 1 (Very much against) to 5 (Very much in favor). *P* values were calculated by ordinary least squares regression with robust standard errors to account for clustering within institution. Disciplines with the same superscript letter are statistically significantly different ( $P < .05$ after Bonferroni adjustment).
\*Responses of “No opinion” are presented for completeness but were not included in this analysis or the sample size column.

Respondents generally reported more positive attitudes for themselves compared to their estimates of the open science attitudes for other faculty within their discipline (Figure 1). Faculty estimated that other faculty within their discipline had the most positive attitudes towards open access publishing (ranging from 42% for ESAS faculty to 57% for clinical and translational sciences faculty). With the exception of code sharing for clinical and translational sciences faculty (41%), estimates of favorable attitudes were below 40% for all other open science practices across pharmacy disciplines. Faculty generally estimated that their colleagues held less favorable attitudes towards open science practices. The differences between their own attitudes versus estimated favorable attitudes ranged from -37.1 percentage points for sharing code in clinical and translational sciences faculty (i.e., respondents estimated that their own attitudes were more favorable than their colleagues) to +2.1 percentage points for posting preprints in clinical and translational sciences faculty (i.e., respondents estimated that their colleagues’ attitudes were more favorable than their own attitudes).

**Figure 1:**
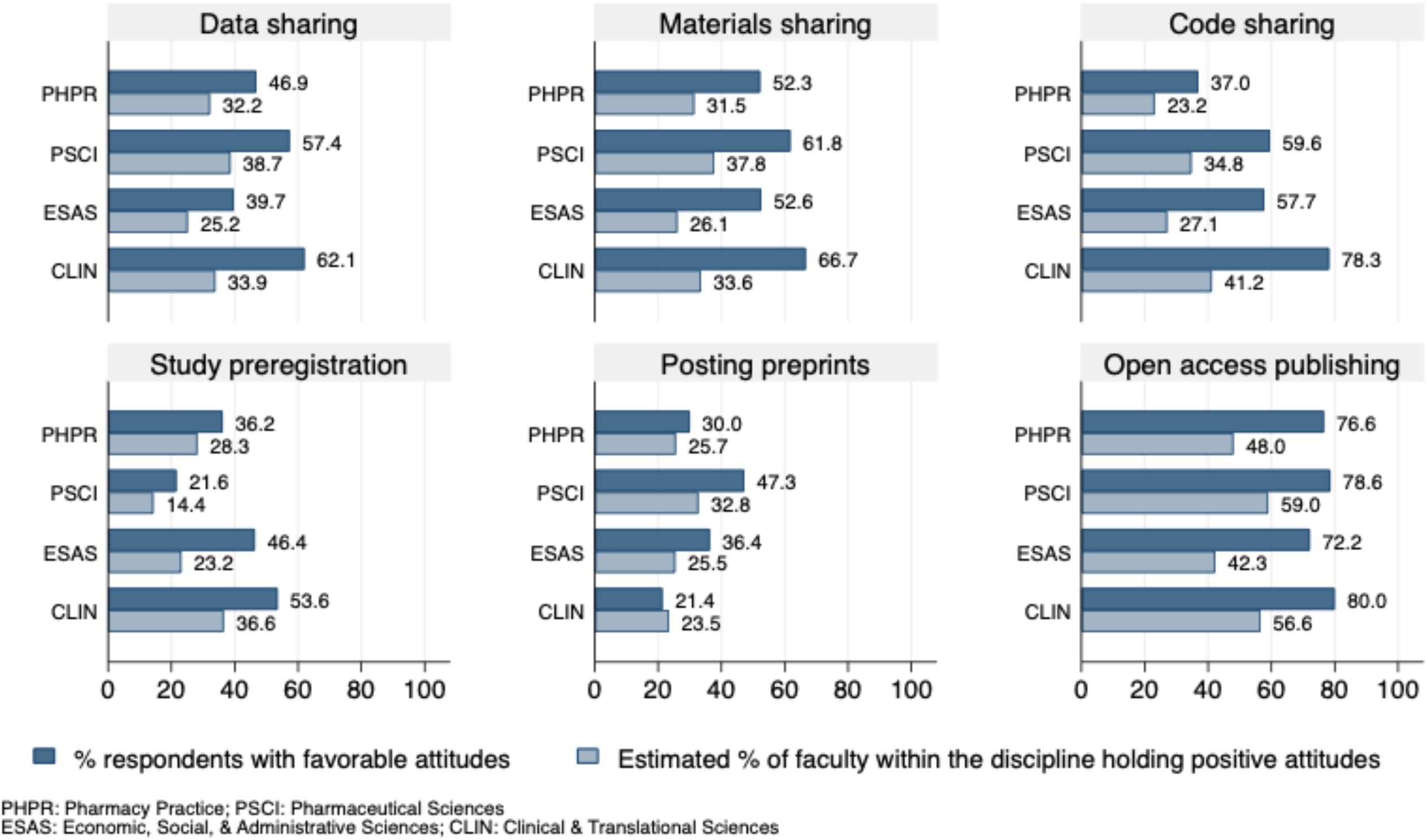
Comparison of Respondents’ Favorable Attitudes and Estimated Favorable Attitudes by Pharmacy Discipline

### 3.5. Open Science Behaviors

Respondents reported less than half of their prior scholarly works had involved open science practices (Figure 2). The most common open science practice was open access publishing (overall mean [SD], 27.7% [29.1%]), which ranged from 23.2% (27.9%) of works for pharmacy practice faculty to 37.5% (30.1%) for clinical and translational sciences faculty. The least common open science practice was study preregistration (overall mean [SD], 1.7% [7.0%]). Aside from open access publishing, there were only 3 instances where the reported percentage of scholarly works was below 10%: data sharing among pharmaceutical sciences faculty, materials sharing among pharmaceutical sciences faculty, and data sharing among clinical and translational sciences faculty.

**Figure 2:**
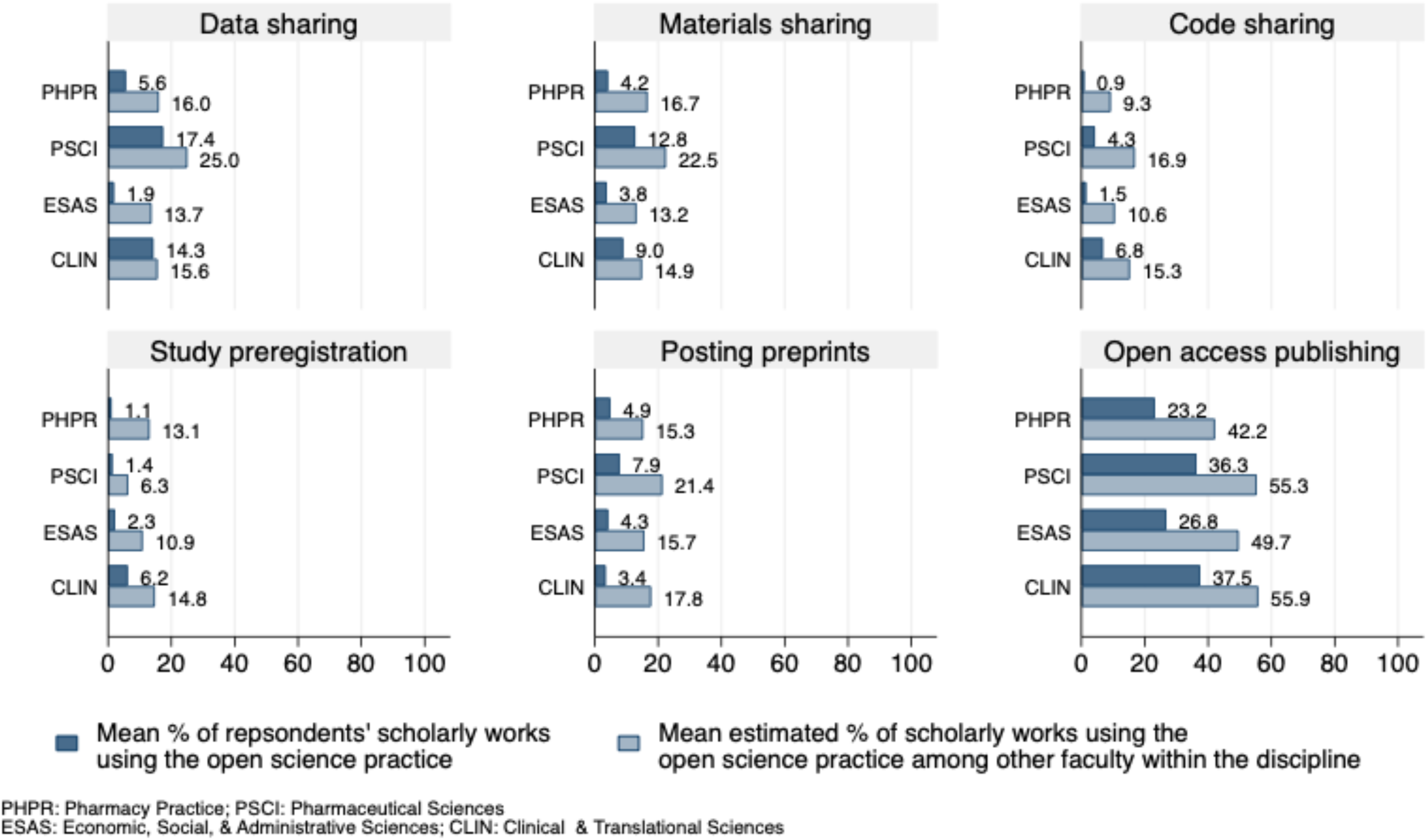
Comparison of Respondents’ Open Science Practices and Estimated Open Sciences Practices by Pharmacy Discipline

The comparison of open science behaviors across pharmacy disciplines is presented in Table 3. For these generalized linear models, pharmacy practice faculty were chosen as the reference group since they were the most common group of respondents and because that group was assumed to be somewhat different from the other disciplines given their primary focus on practice rather than research. From Model 1 (pharmacy discipline as the only predictor), the odds of pharmaceutical sciences or clinical and translational sciences faculty engaging in data sharing, code sharing, or open access publishing were from 2 to almost 8 times higher than pharmacy practice faculty. ESAS faculty had an almost 70% lower odds of sharing data compared to pharmacy practice faculty. Additionally, pharmaceutical sciences faculty had over 3 times higher odds of sharing materials than pharmacy practice faculty and clinical and translational sciences faculty had an almost 6 times greater odds of preregistering studies compared to pharmacy practice faculty.

**Table 3.** Participation in Open Science Practices by Pharmacy Discipline

|  | Data sharing<br>OR (95% CI) | Materials sharing<br>OR (95% CI) | Code sharing<br>OR (95% CI) | Study<br>preregistration<br>OR (95% CI) | Posting preprints<br>OR (95% CI) | Open access<br>publishing<br>OR (95% CI) |
| --- | --- | --- | --- | --- | --- | --- |
| *Model 1: Pharmacy discipline* |  |  |  |  |  |  |
|  | n = 462 | n = 456 | n = 429 | n = 444 | n = 458 | n = 466 |
| Pharmacy practice | — | — | — | — | — | — |
| Pharmaceutical sciences | 3.52 (1.97-6.28)* | 3.35 (1.80-6.23)* | 4.78 (1.31-17.42)* | 1.25 (0.34-4.60) | 1.65 (0.85-3.21) | 1.89 (1.35-2.64)* |
| ESAS | 0.32 (0.15-0.69)* | 0.91 (0.41-2.02) | 1.67 (0.46-6.14) | 2.04 (0.90-4.65) | 0.86 (0.38-1.90) | 1.21 (0.83-1.78) |
| Clinical and translational sciences | 2.78 (1.31-5.92)* | 2.26 (0.96-5.34) | 7.81 (2.06-29.64)* | 5.87 (1.66-20.73)* | 0.69 (0.27-1.74) | 1.99 (1.24-3.19)* |
| *Model 2: Model 1 + gender, racial or ethnic minority, current enrollment in a training program, possessing a research-based doctoral degree, percent effort allocated to research, academic rank, and tenure status* |  |  |  |  |  |  |
|  | n = 308 | n = 304 | n = 291 | n = 297 | n = 307 | n = 309 |
| Pharmacy practice | — | — | — | — | — | — |
| Pharmaceutical sciences | 4.67 (1.12-19.51)* | 2.46 (0.68-8.93) | 1.02 (0.22-4.73) | 2.02 (0.22-18.17) | 0.66 (0.19-2.22) | 2.15 (1.05-4.38)* |
| ESAS | 0.22 (0.06-0.87)* | 0.93 (0.26-3.30) | 0.16 (0.03-0.76)* | 2.28 (0.61-8.46) | 0.36 (0.14-0.95)* | 1.35 (0.66-2.77) |
| Clinical and translational sciences | 1.87 (0.35-9.91) | 1.60 (0.43-5.91) | 1.43 (0.28-7.25) | 6.97 (0.78-62.50) | 0.22 (0.08-0.66)* | 1.63 (0.84-3.18) |
| *Model 3: Model 2 + Carnegie classification for the parent institution and three-year total funding for the school or college of pharmacy* |  |  |  |  |  |  |
|  | n = 253 | n = 251 | n = 241 | n = 246 | n = 254 | n = 255 |
| Pharmacy practice | — | — | — | — | — | — |
| Pharmaceutical sciences | 12.30 (3.04-49.72)* | 2.87 (0.65-12.72) | 2.52 (0.27-23.27) | 5.15 (0.52-51.34) | 1.05 (0.24-4.52) | 2.37 (0.97-5.77) |
| ESAS | 0.32 (0.09-1.16) | 1.41 (0.29-6.86) | 0.28 (0.04-1.77) | 5.20 (1.22-22.24)* | 0.49 (0.16-1.49) | 1.37 (0.58-3.26) |
| Clinical and translational sciences | 3.18 (0.97-10.39) | 1.72 (0.45-6.53) | 2.93 (0.51-16.95) | 21.50 (2.10-220.14)* | 0.31 (0.10-0.97)* | 1.56 (0.74-3.28) |

Abbreviations: OR, odds ratio; CI, Confidence interval; ESAS, Economic, social, and administrative sciences.
All models were estimated using a generalized linear model with the proportion of scholarly works involving the open science activity as the dependent variable using a logit link, binomial distribution, and robust standard errors clustered on institution. Pharmacy practice was the reference group.
\* $P < .05$

In Model 2, gender, identifying as a racial or ethnic minority, current enrollment in a training program, possessing a research-based doctoral degree, percent effort allocated to research, academic rank, and tenure status were included as confounders based on results from previous studies^18,30–34^ and conceptual relevance. Other combinations of individual characteristics were considered but resulted in adverse effects on model performance. The findings from Model 2 showed similar patterns to Model 1.

Three-year total funding for the school or college of pharmacy and a collapsed Carnegie classification were included in Model 3. Each institution’s basic Carnegie classification^35^ was collapsed in the following manner: baccalaureate institution (arts and science focus or diverse field), master’s institution (M1, M2, or M3), doctoral professional university, medical or health sciences university (Special Focus- Medical Schools and Centers, Special Focus-Other Health Professions Schools), and research-intensive universities (R1, R2, or Special Focus-Research Institution). Other combinations of institutional characteristics were considered but resulted in statistical challenges with model estimation. The results from Model 3 were somewhat different than for Models 1 or 2. The odds of data sharing among pharmaceutical sciences faculty were over 12 times higher than for pharmacy practice faculty (OR [95% CI]: 12.30 [3.04–49.72]). ESAS and clinical and translational sciences faculty had a higher odds of study preregistration compared to pharmacy practice faculty (OR [95% CI]: 5.2 [1.22–22.24] and 21.50 [2.10– 220.14], respectively). Clinical and translational sciences faculty had 70% lower odds of posting preprints (OR [95% CI]: 0.31 [0.10–0.97]) than pharmacy practice faculty. There were no statistically significant differences by pharmacy discipline for materials sharing, code sharing, or open access publishing in Model 3. It is important to note the wide confidence intervals for Model 3 suggesting imprecise estimates potentially associated with small sample sizes resulting from additional variables being added to the models.

Figure 2 provides a comparison of the percentage of respondents’ scholarly works using various open science practices with their estimates of the percentage of faculty within their discipline who have used open science practices. Compared to their own scholarly work, respondents estimated that more faculty within their discipline had engaged in open science practices. This ranged from a 1.3 percentage point difference for data sharing in clinical and translational sciences faculty to a 22.9 percentage point difference for open access publishing in ESAS faculty.

## 4. Discussion

This study provides an overview of the current attitudes and behaviors of US pharmacy faculty in relation to selected open science practices. Faculty had the most positive views of open access publishing. This was also the most frequently reported open science practice. This was expected given the rise in popularity and uptake of open access publishing pathways. Although sharing preprints and study preregistration had the least positive responses for attitudes, these were still in the neutral range. This is similar to findings from a 2020 study by Soderberg, Errington, and Nosek^20^ where medical researchers reported the least positive views towards preprints (51% favorable views for medical and health sciences vs. ≥70% favorable views for life, physical, and psychosocial science). This low favorability and slow uptake of preprints in health sciences may arise, in part, from their questionable value and appropriateness.^36–40^ The most frequent open science activity was open access publishing. All other open science practices were relatively infrequent for all pharmacy disciplines. Across pharmacy disciplines, there was notable variability in open science attitudes and behaviors.

The comparisons between respondent attitudes and behaviors with their perceived disciplinary norms revealed an interesting pattern. Respondents rated themselves as having more positive attitudes than the rest of faculty within their discipline. Conversely, respondents believed that more faculty in their discipline engaged in each open science practice than they did in their own research. Whether this reflects a lack of familiarity with the various open science practices or faculty feeling like they are less like their peers than they may be remains unclear and should be examined more closely in the future.

The random sampling strategy used in current study should allow these findings to be generalizable to the broader population of full-tine US pharmacy faculty. The representativeness of the respondents in relation to the known characteristics of US pharmacy faculty and the lack of concern surrounding non-response bias further supports generalizability. There are, however, several limitations to note when interpreting the results of this study. Although there were no major indications of non-response bias or concerns related to sample representativeness, there were some differences as noted previously. The over- representation of economic, social, and administrative sciences faculty among respondents is likely related to this being the author’s discipline. The “name recognition” may have promoted increased responses among ESAS faculty. The proportion of respondents from the clinical and translational sciences was smaller than the other disciplines. It is unclear whether this is an artefact of that being the smallest group of pharmacy faculty or if there were other reasons. The extent to which this may have influenced the results is unknown, but it could have resulted in difficulties identifying differences across groups. In the originally registered study plan, the analysis of attitudes involved ordinal logistic regression. For some open science practices, the proportional odds assumption was violated requiring slightly different model specifications across the open science practices. Analyzing the data using OLS regression indicated similar patterns across pharmacy disciplines with respect to statistical significance and the directions of the relationships. For the sake of consistency, to facilitate interpretation, and to avoid confusion, the results for open science attitudes based on OLS regression were reported. Robust standard errors were used in all models to account for clustering within institutions. Different model specifications could have resulted in different results. The wide confidence intervals for some estimates shown in Table 3 likely reflect small sample sizes, especially for clinical and translational science faculty. Although the directions of the relations were consistent for most open science practices when moving from Model 1 to Model 3, those results should be interpreted with caution given the low frequency of the outcomes.

Research is ongoing with the data collected in this study. This includes an examination of barriers and incentives to researcher engagement in open science practices. Using behavioral models, examining faculty intentions to engage in open science practices is also in process. The current study recruited current US pharmacy faculty; however, current graduate students and post-graduate trainees should be considered in future work as they represent the next generation of pharmacy researchers and faculty. Continued research in this area will be important to identify intervention points to improve the awareness and uptake of open science practices in pharmacy. This is especially true as open science requirements becoming increasingly required by funding agencies.^41–43^

## 5. Conclusions

This study provides a current assessment of attitudes towards and engagement in various open science practices among US pharmacy faculty. Overall, faculty had the most positive views of open access publishing with the lowest attitudes related to posting preprints and study preregistration. Open access publishing was the most common open science practice reported by faculty. Given the relatively low frequency with which other open science practices were reported, there is considerable room for improvement in all other open science practices.

## Data Availability

All data produced will available online at the project's Open Science Framework (OSF) page after acceptance of the paper.

https://osf.io/udywr/

## Notes

### Competing Interest Statement

The authors have declared no competing interest.

### Clinical Protocols

https://osf.io/68rvy

### Funding Statement

This study did not receive any external funding.

### Author Declarations

Institutional Review Board of Midwestern University (Illinois Campus) gave ethical approval for this work (File Number 22011).

